# Pararectus versus ilioinguinal approach for the management of acetabular fractures: A Protocol for Systemic Review and Meta-analysis

**DOI:** 10.1101/2023.04.14.23288547

**Authors:** Rajesh Kumar Rajnish, Amit Srivastava, Sandeep Kumar Yadav, Abhay Elhence, Kuldeep Rathor, Saurabh Gupta

## Abstract

**Background:** Acetabular fractures result from high-energy trauma, and their complex anatomy poses a challenge to surgeons for open reduction and internal fixation of these fractures. The goal of fixation is an anatomical reduction of articular surfaces and stable fixation with minimal damage to vital structures around. The long-term clinical outcomes of these fractures are significantly impacted by selecting an appropriate surgical approach with minimal complications to achieve an anatomical reduction with congruent articular surfaces. The purpose of this review is to compare the outcomes of the pararectus (PR) versus the ilioinguinal (IL) approach for the open reduction and internal fixation (ORIF) of displaced acetabular fractures by looking at the evidence in the existing literature.

**Methods:** A systematic review and meta-analysis will be performed in accordance with the PRISMA guidelines. A primary search will be done through the electronic databases of PubMed/Medline, Embase, Scopus, and the Cochrane Library using a pre-defined search strategy. A comparative study, either randomized control trials (RCTs) or non-randomized trials, which have compared at least one outcome of the internal fixation of acetabular fractures using the PR versus IL approach will be included in the current review.

Single-armed non-comparative studies, conference posters, abstracts, case reports, review articles, cadaveric studies, book chapters, technical tips, and biomechanical studies will be excluded. Both qualitative and quantitative data analysis will be performed. Appropriate tables and diagrams will be used to demonstrate the qualitative data presentation, and wherever feasible, a quantitative analysis will be done with the appropriate software. The risk-of-bias assessment for non-randomized comparative studies will be done using the MINORS tool, and the Cochrane Collaborations risk-of-bias tool will be used for RCTs.

## 1. Background

Acetabular has a complex anatomy, surrounded by a bulk of soft tissues and neurovascular structures. Most commonly in young adults these fractures result from high-energy trauma [1]. Its deep, complex anatomy poses a significant challenge to surgeons for ORIF of these fractures. The goal of surgical fixation is an anatomical reduction and stable fixation with minimal soft tissue damage and complications [1,2,3]. Hence, choosing an appropriate surgical approach wisely is of great importance in achieving an anatomical reduction with optimal long-term outcomes. Therefore, an appropriate surgical approach must be chosen according to the fracture pattern and the anticipated complications. [4]

The IL approach is considered a classical approach for acetabular fracture fixation, which provides good exposure to the fracture site. This approach is commonly used for the fixation of anterior acetabular fractures, allowing indirect access to the quadrilateral surface. [5,6] In addition to these advantages, the IL approach too associated with disadvantages, such that it is an extensile approach with a long incision length and requires three windows to access fracture. It has a longer duration of surgery and is associated with a few serious complications like neurovascular injury, vasospasm, thrombus formation, and inguinal hernia because of complex local anatomy [1,3,6].

The PR approach, a new surgical approach introduced by Keel et al. [7] for anterior acetabular fracture fixation, is a single-incision approach, less invasive, and allows direct access to the iliac fossa, quadrilateral plate, acetabular dome, and pubic symphysis. The additional advantage of this approach is that it combines the intrapelvic access offered by the second window of the modified Stoppa approach and the third window of the IL approach with fewer complications [1,7].

However, this is still a question of debate to establish the superiority and safety of the PR approach over the IL approach for acetabular fracture fixation. Hence, this systematic review and meta-analysis were planned to compare the perioperative and postoperative outcomes and complications of the PR and IL approach.

## 2. Need for review

Although the PR approach emerged as a versatile approach that provides greater access to the false pelvis and the posterior pelvic ring without additional surgical window and patient position change. However, the effectiveness and safety of the PR approach over the IL approach have still to be widely accepted for acetabular fracture fixation. To the best of my knowledge, only one published meta-analysis has compared the PR approach to the other surgical approach for acetabular fracture with limited evidence. Hence, this systematic review and meta-analysis compare the perioperative and postoperative outcomes and complications of the PR and IL approach.

## 3. Objectives

### Primary Objectives

i. To compare the primary outcomes reduction quality, complications of internal fixation of acetabular fracture through pararectus versus ilioinguinal approach.

### Secondary Objective

i. Additionally, to compare the duration of surgery, intraoperative blood loss, and surgical incision length between two surgical approaches.

## 4. PICO framework for the study

Participants: acetabulum fracture

Intervention: Pararectus approach

Control: Ilioinguinal approach

Outcomes: Reduction quality, complications, duration of surgery, blood loss, and surgical incision length.

## 5. Methods

i. Review Protocol: This systematic review and meta-analysis will be performed in accordance with the Preferred Reporting Items for Systematic Reviews and Meta-analysis (PRISMA) guidelines. [8] A protocol for the review will be formulated in accordance with the PRISMA-P guidelines. (Appendix I)
ii. Eligibility Criteria: A comparative study, either randomized control trials (RCTs) or non-randomized, will be included, which compares the outcomes of the internal fixation of PR versus IL approach used for ORIF of acetabular fractures and report at least one primary or secondary outcome of the review. Single-arm studies that do not compare the outcomes of the internal fixation of PR versus IL approach, case reports, conference abstracts, posters, book chapters, review articles, cadaveric studies, biomechanical studies, and technical tips, will be excluded.
iii. Information Sources & Literature search: A primary literature search will be conducted on the Medline, Embase, Scopus, and Cochrane Library databases, using pre-defined search strings of keywords *“ ((acetabular fractures*) AND (pararectus approach)) AND (ilioinguinal approach)”* A manual secondary search of the bibliography of the full text of all included articles and relevant review articles will be conducted.
iv. Study Selection: All the identified articles will be screened through titles and abstracts for eligibility independently by three authors. After initial screening, full texts of all selected articles will be obtained. The reasons for excluding those articles for which full text was obtained will be documented. Any discrepancies in the article selection process will be resolved by mutual agreement between the authors.
v. Data Collection & Data Items: Data will be extracted on pre-formed data collection forms by two authors independently, and a third author will cross-check the data for accuracy. Baseline data items will include:

1. Name of authors and year of publication
2. Number of patients/cases
3. Study design
4. Type of approach used
5. Number of patients in each group
6. Mean age
7. Gender ratio
8. Fracture classification
9. Mean follow up
10. Primary and Secondary Outcomes of the Study

vi) Outcome Measures: The following outcome measures will be evaluated; however, addition or modifications will be made if needed: the primary outcomes of interest will be reduction quality, overall complication, neuro-vascular injury, infection, deep vein thrombosis, heterotopic ossification, and pulmonary embolism. The secondary outcomes of interest will be the duration of surgery, intraoperative blood loss, and surgical incision length.
vii) Data Analysis and Synthesis: A qualitative data synthesis will be performed with appropriate tables and data visualization diagrams. The quantitative synthesis will be performed if ≥ 2 studies included in this review reported the values of either the primary or secondary outcomes of interest. To describe the measure of treatment effects, the mean difference will be used for continuous variables, and the odds ratio will be used for dichotomous variables. All the results will be expressed along with 95% confidence intervals. Forest plots will be made to visualize the results in diagrammatic representation. The statistical heterogeneity will be determined by using the I-square test. Reasons for clinical heterogeneity, if any, will be explored. If the heterogeneity is low (I-square value near 0%) fixed-effects model otherwise, the random effects model will be used. If possible, subgroup analysis will be performed. Publication bias will be estimated and will be shown with a funnel plot using one of the primary outcomes. Meta-analysis will be performed by using Review Manager Software version 5.4. [9]
viii) Assessment of Risk of Bias: The risk-of-bias assessment will be done using the MINORS tool for the non-RCTs [10], and the Cochrane Collaboration’s risk-of-bias tool [11] will be used for RCTs.

## Data Availability

Yes

---

**APPENDIX 1.** PRISMA-P (Preferred Reporting Items for Systematic review and Meta-Analysis Protocols) 2015 checklist: recommended items to address in a systematic review protocol* **\* It is strongly recommended that this checklist be read in conjunction with the PRISMA-P Explanation and Elaboration (cite when available) for important clarification on the items. Amendments to a review protocol should be tracked and dated. The copyright for PRISMA-P (including checklist) is held by the PRISMA-P Group and is distributed under a Creative Commons Attribution Licence 4.0**.

| Section and topic | Item No | Checklist item | Page Number |
| --- | --- | --- | --- |
| **ADMINISTRATIVE INFORMATION** |  |  |  |
| Title: |  | Pararectus versus ilioinguinal approach for the management of acetabular fractures: A Protocol for Systemic Review and Meta-analysis | 1,2 |
| Identification | 1a | Identify the report as a protocol of a systematic review | 1,2 |
| Update | 1b | If the protocol is for an update of a previous systematic review, identify as such | NA |
| Registration | 2 | If registered, provide the name of the registry (such as PROSPERO) and registration number | None |
| Authors: |  | Rajesh Kumar Rajnish, Amit Srivastava, Sandeep Kumar Yadav, Kuldeep Rathor, Abhay Elhence, Saurabh Gupta | 1 |
| Contact | 3a | Provide name, institutional affiliation, email address of all protocol authors; provide physical mailing address of corresponding author | 1 |
| Contributions | 3b | Describe contributions of protocol authors and identify the guarantor of the review | 1 |
| Amendments | 4 | If the protocol represents an amendment of a previously completed or published protocol, identify as such and list changes; otherwise, state plan for documenting important protocol amendments | NA |
| Support: |  |  |  |
| Sources | 5a | Indicate sources of financial or other support for the review | 1 |
| Sponsor | 5b | Provide name for the review funder and/or sponsor | 1 |
| Role of sponsor or funder | 5c | Describe roles of funder(s), sponsor(s), and/or institution(s), if any, in developing the protocol | None |
| **INTRODUCTION** |  |  |  |
| Rationale | 6 | Describe the rationale for the review in the context of what is already known | 3 |
| Objectives | 7 | Provide an explicit statement of the question(s) the review will address with reference to participants, interventions, comparators, and outcomes (PICO) | 4 |
| **METHODS** |  |  |  |
| Eligibility criteria | 8 | Specify the study characteristics (such as PICO, study design, setting, time frame) and report characteristics (such as years considered, language, publication status) to be used as criteria for eligibility for the review | 4,5 |
| Information sources | 9 | Describe all intended information sources (such as electronic databases, contact with study authors, trial registers or other grey literature sources) with planned dates of coverage | 5 |
| Search strategy | 10 | Present draft of search strategy to be used for at least one electronic database, including planned limits, such that it could be repeated | 5 |
| Study records: |  |  |  |
| Data management | 11a | Describe the mechanism(s) that will be used to manage records and data throughout the review | 5 |
| Selection process | 11b | State the process that will be used for selecting studies (such as two independent reviewers) through each phase of the review (that is, screening, eligibility and inclusion in meta-analysis) | 5 |
| Data collection process | 11c | Describe planned method of extracting data from reports (such as piloting forms, done independently, in duplicate), any processes for obtaining and confirming data from investigators | 5 |
| Data items | 12 | List and define all variables for which data will be sought (such as PICO items, funding sources), any pre-planned data assumptions and simplifications | 5 |
| Outcomes and prioritization | 13 | List and define all outcomes for which data will be sought, including prioritization of main and additional outcomes, with rationale | 5 |
| Risk of bias in individual studies | 14 | Describe anticipated methods for assessing risk of bias of individual studies, including whether this will be done at the outcome or study level, or both; state how this information will be used in data synthesis | 6 |
| Data synthesis | 15a | Describe criteria under which study data will be quantitatively synthesised | 6 |
| | 15b | If data are appropriate for quantitative synthesis, describe planned summary measures, methods of handling data and methods of combining data from studies, including any planned exploration of consistency (such as $I^2$, | 6 |
| | | Kendall's $\tau$) | |
|  | 15c | Describe any proposed additional analyses (such as sensitivity or subgroup analyses, meta-regression) | 6 |
|  | 15d | If quantitative synthesis is not appropriate, describe the type of summary planned | 6 |
| Meta-bias(es) | 16 | Specify any planned assessment of meta-bias(es) (such as publication bias across studies, selective reporting within studies) | 6 |
| Confidence in cumulative evidence | 17 | Describe how the strength of the body of evidence will be assessed (such as GRADE) | Not Planned |

*From: Shamseer L, Moher D, Clarke M, Ghersi D, Liberati A, Petticrew M, Shekelle P, Stewart L, PRISMA-P Group. Preferred reporting items for systematic review and meta-analysis protocols (PRISMAP) 2015: elaboration and explanation. BMJ. 2015 Jan 2;349(jan02 1):g7647.*

## References

1. Zou R, Wu M, Guan J, Xiao Y, Chen X. Clinical Results of Acetabular Fracture via the Pararectus versus Ilioinguinal Approach. Orthop Surg. 2021 Jun;13(4):1191–1195.

2. Märdian S, Schaser KD, Hinz P, Wittenberg S, Haas NP, Schwabe P. Fixation of acetabular fractures via the ilioinguinal versus pararectus approach: a direct comparison. Bone Joint J. 2015 Sep;97-B(9):1271–8.

3. Srivastava A, Rajnish RK, Kumar P, Haq RU, Dhammi IK. Ilioinguinal versus modified Stoppa approach for open reduction and internal fixation of displaced acetabular fractures: a systematic review and meta-analysis of 717 patients across ten studies. Arch Orthop Trauma Surg. 2023 Feb;143(2):895–907.

4. Letournel E. The treatment of acetabular fractures through the ilioinguinal approach. Clin Orthop Relat Res.1993; 292:62–76.

5. Letournel E. Fractures of the acetabulum. A study of a series of 75 cases. 1961. Clin Orthop Relat Res.1994;305:5

6. Matta, Joel M MD. Operative Treatment of Acetabular Fractures Through the Ilioinguinal Approach: A 10-Year Perspective. Journal of Orthopaedic Trauma. 2006; 20(1):p S20–S29.

7. Keel MJ, Ecker TM, Cullmann JL, Bergmann M, Bonel HM, Büchler L, Siebenrock KA, Bastian JD. The Pararectus approach for anterior intrapelvic management of acetabular fractures: an anatomical study and clinical evaluation. J Bone Joint Surg Br. 2012 Mar;94(3):405–11.

8. Page MJ, Moher D, Bossuyt PM, et al. PRISMA 2020 explanation and elaboration: updated guidance and exemplars for reporting systematic reviews. BMJ. 2021 Mar 29;72:160.

9. Review Manager (RevMan) [Computer program]. Version 5.4, The Cochrane Collaboration, 2020.

10. Slim K, Nini E, Forestier D, Kwiatkowski F, Panis Y, Chipponi J. Methodological index for nonrandomized studies (minors): development and validation of a new instrument. ANZ J Surg.2003; 73(9):712–6.

11. Higgins JPT, Altman DG, Gotzsche PC, Juni P, Moher D, Oxman AD, et al. Cochrane Bias Methods Group Cochrane Statistical Methods Group. The Cochrane Collaboration’s tool for assessing the risk of bias in randomized trials. BMJ 2011; 343:d5928.

